# Ultrasensitive assay for saliva-based SARS-CoV-2 antigen detection

**DOI:** 10.1101/2021.02.17.21251863

**Authors:** A Ren, D Sohaei, I Zacharioudakis, GB Sigal, M Stengelin, A Mathew, C Campbell, N Padmanabhan, D Romero, J Joe, A Soosaipillai, V Kulasingam, T Mazzulli, XA Li, A McGeer, EP Diamandis, I Prassas

## Abstract

Widespread SARS-CoV-2 testing is highly valuable for identifying asymptomatic/pre-symptomatic individuals to slow community disease transmission. However, there remains a technological gap for highly reliable, easy, and quick SARS-CoV-2 diagnostic tests that are suitable for frequent mass testing. Compared to the conventional nasopharyngeal (NP) swab-based tests, saliva-based methods are attractive due to easier and safer sampling protocols. Despite its merits in rapid turn-around-time and high throughput compared to traditional PCR-based technologies, the widespread use of saliva-based SARS-CoV-2 rapid antigen tests is hindered by limited analytical sensitivity of current methods. Here, we report the first ultrasensitive, saliva-based SARS-CoV-2 antigen assay with an analytical sensitivity of < 0.32 pg/ml, corresponding to 4 viral RNA copies/µl, which is comparable to that of PCR-based tests. Using the novel electrochemiluminescence (ECL)-based S-PLEX immunoassay, we measured the SARS-CoV-2 nucleocapsid (N) antigen concentration in 105 saliva samples obtained from non-COVID-19 and COVID-19 patients. Our assay displayed absolute specificity and high sensitivity (90.2%), where it correctly identified samples with viral loads up to 35 CT cycles by saliva-based PCR. Paired NP swab-based PCR results were also obtained for 86 cases for comparison. Our assay showed high concordance with saliva-based and NP swab-based PCR in samples with negative (< 0.32 pg/ml) and strongly positive (> 2 pg/ml) N antigen concentrations. Our study unveiled the ultrasensitivity and specificity of the saliva-based S-PLEX assay, demonstrating its clinical value as a high throughput, complementary alternative to PCR-based techniques. The novel technique is especially valuable in cases where compliance to frequent swabbing may be problematic (e.g. schools, nursing homes, etc.).

## Introduction

Despite the approval of the first vaccines against SARS-CoV-2, the COVID-19 pandemic remains a significant global threat and the return to a pre-pandemic ‘normalcy’ is still projected to be long and turbulent ^1^. Considering the significant isolation fatigue and the diminishing tolerance for horizontal lockdowns, we urgently need better strategies for preventing disease spread until significant vaccine-induced herd immunity is achieved. Frequent SARS-CoV-2 population testing, in combination with isolation and contact tracing, have been demonstrated to be effective for slowing transmission ^2^. Mass testing strategies depend on reliable, easy, and quick SARS-CoV-2 diagnostic tests, which could be used routinely and frequently to identify asymptomatic/pre-symptomatic individuals in both healthcare and non-healthcare settings (e.g. nursing homes, large enterprises, institutions/schools, etc.) ^3^.

While nasopharyngeal (NP) swab-based testing has been primarily used as the gold standard method for SARS-CoV-2 diagnosis, saliva-based technologies have emerged as a promising alternative with excellent fit for mass testing applications ^4–7^. Unlike NP swabbing, saliva sampling is easy, non-invasive, painless, and safe, by eliminating the need for close contact between patients and medical personnel during sample collection and the associated risk of disease transmission. Several PCR-based studies have compared saliva-versus nasal (or nasopharyngeal/oropharyngeal) swab-based SARS-CoV-2 nucleic acid detection post-infection, showing an overall similar performance ^8^. Despite their outstanding sensitivity, traditional PCR-based technologies rely on sophisticated equipment and trained personnel, factors that collectively limit their applicability for mass-oriented testing campaigns. Efforts for more portable nucleic acid-based methods have been recently reported ^9^. Additionally, PCR-based technologies can be error-prone (due to multiple pre-analytical sample preparation steps), are semi-quantitative, and their performance can be influenced by operational and sampling skills (e.g. during swabbing) ^10^.

A more attractive alternative is the use of saliva for direct SARS-CoV-2 antigen detection. To date, the widespread use of saliva-based SARS-CoV-2 rapid antigen testing has been hampered by the relatively low analytical sensitivity of the lateral flow assays (LFAs) that make up the bulk of the available antigen tests. Typical lower limits of detection (LLD) for SARS-CoV-2 LFAs exceed 100 pg/ml ^11^. When applied to saliva, these tests are suitable for detecting cases with high viral loads (e.g. more than 100,000 copies/ml). However, their reliability is significantly compromised in cases with lower but clinically significant loads ^12^.

Here, we report the first ultrasensitive, saliva-based, N antigen assay with potential mass screening applications (with a high throughput of > 2,000 tests per machine per 8 hours). Using a novel electrochemiluminescence (ECL)-based immunoassay (from now on referred to as S-PLEX assay), we demonstrate the ultrasensitive SARS-COV-2 nucleocapsid (N) antigen detection in saliva. The analytical sensitivity at our assay threshold (< 0.32 pg/ml), corresponds to 4 viral RNA copies/µl ^11^, on par with the sensitivity of traditional PCR-based molecular tests. Using this quantitative and sensitive technique we also characterized the range of antigen concentrations found in the saliva of COVID-19 patients, generating information that will be critical in establishing the required performance characteristics for future saliva-based rapid antigen tests.

## Study Design

### Clinical Samples

A total of 105 retrospectively collected saliva specimens were obtained under approval of the Sinai Health System Research Ethics Board (REB#: 02-0118U). Fifteen samples were collected from non-COVID-19 patients (no prior COVID-19 diagnosis) and 90 from patients who were diagnosed with COVID-19 by clinical NP swab or midturbinate nasal swab-based PCR at a network of hospitals including Sinai Health System, Sunnybrook Health Centre, North York General Hospital, and Michael Garron General Hospital (Toronto, Canada). Samples were collected at different time points post-symptom onset (ranging from 1–42 days) or at other disease diagnosis, for the non-COVID-19 samples. **Supplemental Table 1** summarizes the entire saliva cohort used in this study. Paired NP swab sampling was also performed on the day of saliva collection for 86 samples, for comparison purposes. For saliva samples, patients were asked under informed consent to spit into a sterile 50-mL specimen container which was topped with 2.5 ml of phosphate-buffered saline. Samples were transported to the microbiology laboratory at Sinai Health System, where they were treated with 1% Triton X 100 at room temperature for 1 hour before being frozen at −80°C, within 8 hours from time of collection, using a standardized protocol. Prior to analysis, all samples were subsequently thermally inactivated by incubation at 65 °C for 30 min.

### Saliva/swab-based PCR and saliva-based antigen testing using S-PLEX assay

The saliva and swab samples were extracted with the QIAamp Viral RNA Mini Kit (Qiagen). The extracted samples were tested by reverse transcription PCR on the Rotor-Gene Q PCR cycler (Qiagen) using an in-house protocol developed by the Shared Hospital Laboratory ^13^, which targets the E gene and the 5’UTR. All N antigen values were obtained with the S-PLEX ultrasensitive ECL immunoassay platform (Meso Scale Diagnostics) in a fully blinded fashion and according to manufacturer protocols (accessible online at https://www.mesoscale.com/en/products/S-PLEX-SARS-CoV-2-n-kit-sector-1-pl-k150adhs/). All samples were diluted an additional two-fold prior to analysis. As described previously (11), the lower limit of detection (LLD) for the assay, defined as the concentration (uncorrected for sample dilution) that provides a signal 2.5 standard deviations above the assay background signal, was determined to be 0.16 pg/ml. A threshold concentration of 2 x LLD (0.32 pg/ml) was selected as the cut-off for classifying samples as COVID-19 positive or negative.

### Statistical Analysis

All data analysis was performed in GraphPad PRISM 9. Graphs were generated in TIBCO Spotfire Desktop 10.6.1. For statistical analysis of the saliva antigen data, all samples with concentration values (prior to correction for dilution) lower than the LLD of the assay (0.16 pg/ml) were assigned the value of the LLD (0.16 pg/ml). All values higher than the upper limit of detection (1,000 pg/ml) were also assigned the value of the upper limit (1,000 pg/ml). Correlation analysis between saliva antigen concentrations and saliva-based PCR CT values was performed with Spearman correlation test.

Nonparametric Kruskal-Wallis test was performed for group comparisons between saliva antigen concentrations in the following three groups: 1) Saliva-based PCR negative results (non-COVID-19 cases), 2) saliva-based PCR results with CT value ≥ 35 (deemed as indeterminate COVID-19 cases), and 3) saliva-based PCR positive results with CT value < 35 (deemed as potentially active COVID-19 cases). Dunn’s test was performed to correct for multiple comparisons. A P value of < 0.05 was considered statistically significant.

## Results

The LLD of the S-PLEX SARS-CoV-2 N assay was 0.16 pg/ml (0.16 fg/µl) and a threshold value of 2 X LLD (0.32 pg/ml) was used as a cut-off for classifying samples as positive or negative. It has been previously calculated that one viral RNA copy corresponds to 0.15 fg of N antigen, indicating an extremely high analytical sensitivity for the S-PLEX assay of ∼1–2 RNA copies/µl ^11^. This LLD of the S-PLEX assay is on par with the sensitivity of most PCR-based methods and is at least 500 times more sensitive than most commercial LFA-based SARS-CoV-2 antigen assays ^11^. Samples were run in duplicate, where all coefficients of variation (CVs) observed were generally below 10%. Overall, there was a strong negative correlation [Spearman correlation coefficient (r) = −0.864, P < 0.0001] between the N antigen values from S-PLEX and the saliva PCR CT values (**Figure 1**). For group comparisons, the COVID-19 group was further stratified into saliva PCR CT values of ≥ 35 (deemed as PCR negative) to represent indeterminate COVID-19 cases, and saliva PCR CT values of < 35 (deemed as PCR positive) to represent potentially active COVID-19 cases. **Figure 2** shows the distribution of all saliva antigen concentrations (pg/ml) obtained by the S-PLEX assay in the 1) non-COVID-19 PCR-negative group, 2) COVID-19 PCR negative group where CT values were ≥ 35 (considered as indeterminate COVID-19), and 3) COVID-19 PCR positive group where CT values were < 35 (labelled as potentially active COVID-19). Antigen concentration values obtained by S-PLEX ranged between the value of the assay threshold (0.32 pg/ml) and the upper limit of detection (1,000 pg/ml). We further assigned the N concentrations into a weakly positive range (0.33–2.0 pg/ml) and strongly positive range (> 2.0 pg/ml). All 15 non-COVID-19 patients were indeed identified as negatives (below the assay threshold) with the S-PLEX assay (specificity 100%). Among the 64 indeterminate COVID-19 cases, which tested negative using saliva PCR (CT cycles ≥ 35), the S-PLEX assay identified 12 (18.8%) of them as weakly positive (N antigen concentrations ranging from 0.36–1.6 pg/ml). Six of these 12 cases were confirmed to be positive by paired NP swab PCR (CT values < 35), suggesting that these six cases are likely false negatives by the saliva PCR assay. From the 41 potentially active COVID-19 cases, which tested positive using saliva PCR (CT cycles < 35), the S-PLEX assay also identified 37 of them as positive (sensitivity 90.2%). The four cases (patients 6, 20, 71, 75) presumably missed by the S-PLEX test had corresponding saliva PCR CT values of 30, 33, 33, and 31 (**Supplemental Table 1**). Paired NP swab PCR testing was performed for patients 6 and 20 (the other two patients had no swab samples) showing borderline results with CT values of 36 and 33, respectively (**Supplemental Table 1**). The overall concordance of the three assays (S-PLEX saliva antigen, saliva-based PCR, paired NP swab-based PCR) is shown in **Table 1**. As expected, the concordance of the three assays was lowest in the weakly positive range of the S-PLEX assay results (antigen concentrations ranging from 0.33–2 pg/ml) (**Table 1**). Among the 23 saliva samples that tested in this weakly positive range, as determined by the S-PLEX test, 52% of the samples (12/23) tested negative by saliva PCR (≥ 35 CT). Six of these 12 samples were confirmed to be positive by NP swab PCR (< 35 CT), suggesting that the saliva PCR results in these six cases are likely false negatives. The concordance with saliva-based PCR was very high in the S-PLEX assay negative and strongly positive results ranges (92.9% and 100% respectively). Finally, the S-PLEX saliva-based assay and saliva-based PCR showed similar concordance to NP swab PCR.

**Table 1.** Concordance between diagnosis made according to S-PLEX antigen test and PCR-based methods.

| S-PLEX N assay results range | S-PLEX nucleocapsid (N) antigen test concordance with |  | Saliva-based PCR concordance with NP swab-based PCR |
| --- | --- | --- | --- |
|  | Saliva-based PCR* | Nasopharyngeal (NP) swab-based PCR* |  |
| Negative ($\leq 0.32$ pg/ml) | 92.9% | 69.6% | 73.2% |
| Weakly positive (0.33–2.0 pg/ml) | 47.8% | 65.2% | 65.2% |
| Strongly positive ($> 2.0$ pg/ml) | 100.0% | 88.5% | 88.5% |

\*Cut-off for positivity in the saliva- and NP swab-based PCR methods was set to CT cycle value of $< 35$ .
See text for more details

**Figure 1.**
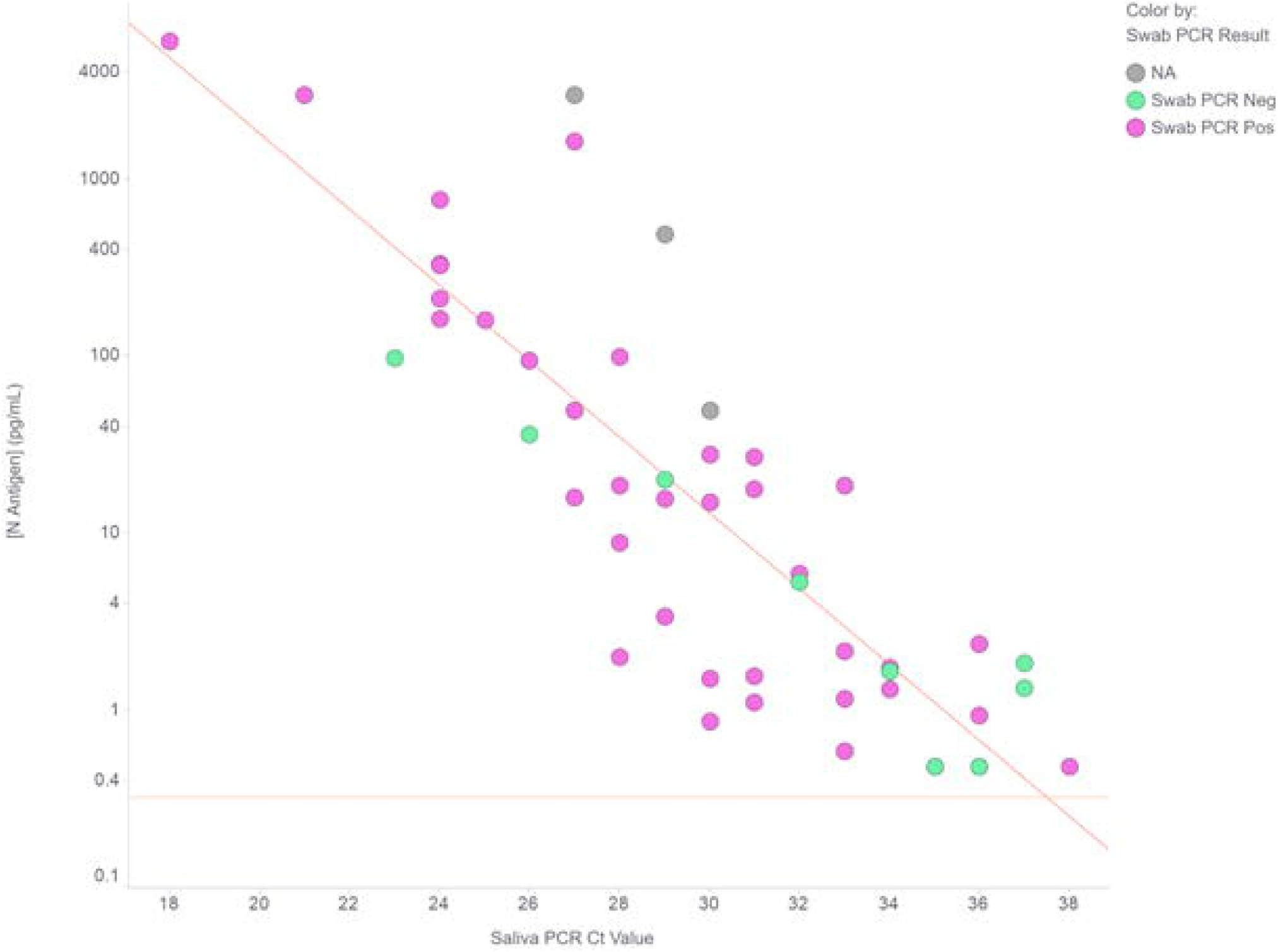
Correlation of viral nucleocapsid protein concentration with PCR CT cycle value for clinical saliva samples from PCR-positive patients. Scatterplot analysis showed strong negative correlation between nucleocapsid (N) concentration (pg/ml) in saliva as determined by the novel ultrasensitive antigen assay with PCR CT cycle value for SARS-CoV-2 RNA in saliva from PCR-positive patients (N = 50) (Spearman coefficient of correlation (r) = −0.864, P < 0.0001). Data points are colored based on the presence or absence of detectable SARS-CoV-2 RNA in paired nasopharyngeal swab samples from the same patient cohort. All N antigen concentrations were corrected for dilution, relative to the original saliva sample, and represent concentrations present in the saliva sample. Horizontal red dotted line indicates the nucleocapsid concentration cut-off of the antigen test for weakly positive samples, which was set at 2 x LLD (0.32 pg/ml). All samples assessed for correlation are PCR-positive.

**Figure 2.**
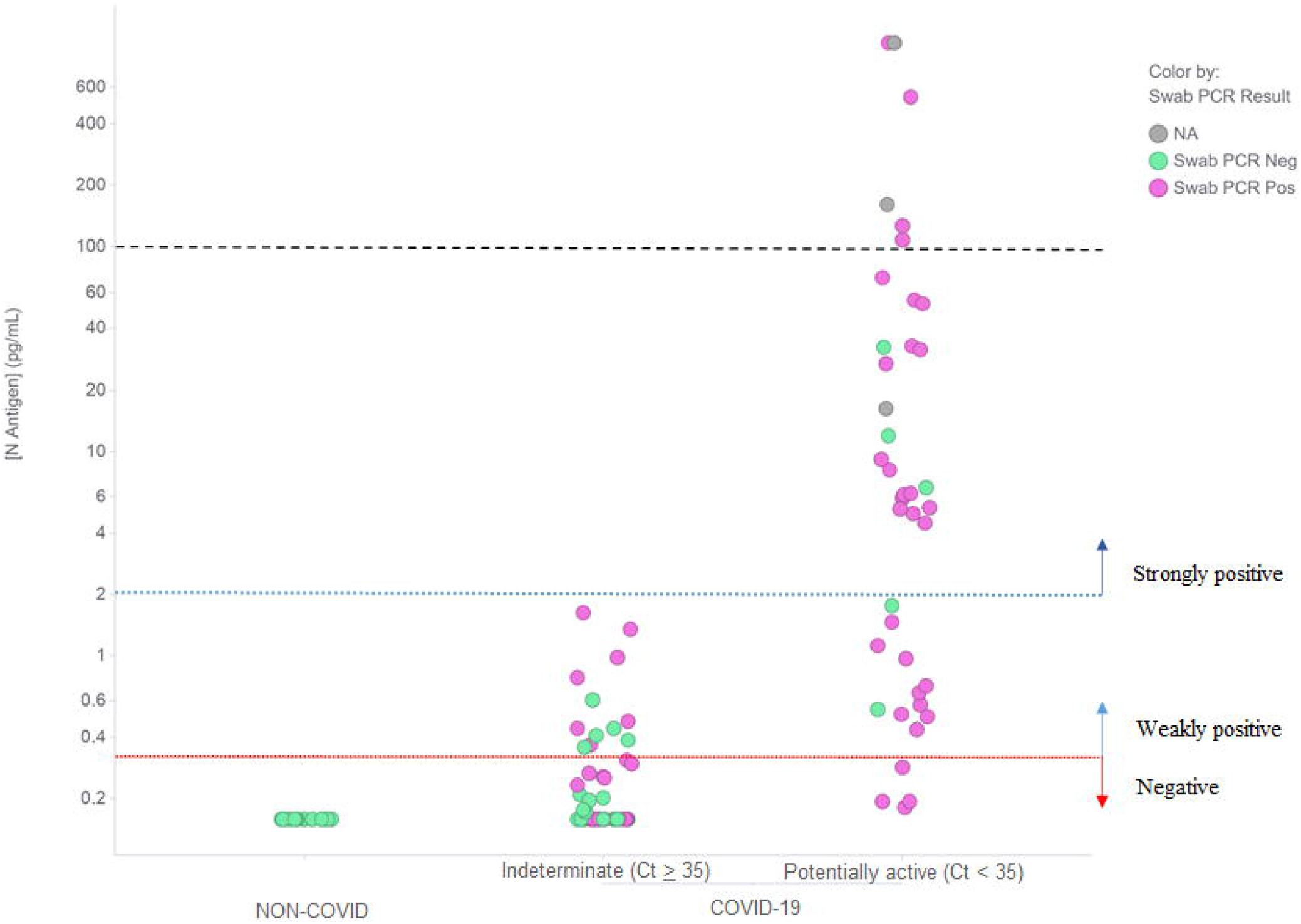
Nucleocapsid concentrations in saliva samples from non-COVID-19 and COVID-19 cases. Saliva-based PCR positivity cut-off in the COVID samples was set at CT cycle value of < 35 to indicate potentially active COVID-19 cases. Nucleocapsid concentration as measured by the novel ultrasensitive antigen assay (uncorrected for dilution) was significantly higher in the saliva PCR-positive samples (saliva-based PCR CT value < 35, potentially active COVID-19 cases) compared to the non-COVID patients and saliva PCR-negative samples (CT value ≥ 35, indeterminate COVID-19 cases) (Kruskal-Wallis test, P < 0.0001). Horizontal red dotted line indicates the nucleocapsid concentration cut-off of the antigen test for weakly positive samples, which was set at 2 x LLD (0.32 pg/ml). The horizontal blue dotted line at 2 pg/ml indicates the cut-off for strongly positive samples. The green dots represent the patients that showed nasopharyngeal swab PCR-positive results (CT value < 35), which suggests their saliva PCR results were false negatives. The pink dots represent the nasopharyngeal swab PCR-negative samples (CT value ≥ 35), while the gray dots represent samples that did not have paired nasopharyngeal swab PCR data. The horizontal black dashed line at 100 pg/ml indicates the typical sensitivity of currently available commercial direct to consumer rapid antigen tests.

## Discussion

To our knowledge, this is the first demonstration of an ultrasensitive immunoassay for SARS-CoV-2 N antigen detection in saliva. Unlike the standard NP swab-based molecular tests, salivary testing is more convenient, safe, non-invasive, and does not require any specialized reagents or technical expertise for proper sampling. Taken together, these factors can warrant an increased compliance in community-based mass screening campaigns. The original notion that NP swabs represent a more reliable sampling source for SARS-CoV-2 detection has been recently challenged by several studies showing a comparable diagnostic sensitivity between saliva-based and NP swab-based PCR tests ^8^. Furthermore, emerging data are now suggesting that viral load estimations, as determined from saliva-based methods, may exhibit a more reliable and dynamic correlation to COVID-19 severity and mortality, compared to NP swab-based modalities ^14^. Of note, posterior oropharyngeal saliva should not be considered equivalent to oral saliva. The first is a part of respiratory secretions, while the second is produced by the salivary glands, which are not part of the respiratory tract.

We expect that saliva-based SARS-CoV-2 testing to become more mainstream in the next phases of the COVID-19 pandemic management. Currently, saliva-based PCR methods are the closest alternative to standard NP swab-based PCR. However, all types of nucleic acid amplification technologies can be impacted by technological caveats, especially during sudden outbreaks in rural areas. Additionally, PCR-technologies are semi-quantitative, and the resulting CT values may vary between instruments, genes, and/or operators. Hence, alternative non-competing quantitative platforms that could complement the existing PCR-based diagnostic capacity should be urgently explored to enhance total testing capacity. Direct to consumer lateral flow rapid antigen tests (RATs) are promising (and as yet largely unexplored) tools for curbing large transmission chains. However, their utility, especially for saliva-based screening, is hampered by their relatively lower sensitivity thresholds ^11^. For instance, in the case of our study presented here, a RAT test with an LLD of 100 pg/ml (typically seen in FDA-approved LFA-based antigen assays) would erroneously miss the majority of the potentially active COVID-19 cases (reducing the sensitivity from 90.2% to 17.1%) (as shown by drawing a horizontal dashed line at a LLD of 100 pg/ml in **Figure 2**).

This is the first in-depth characterization of SARS-CoV-2N antigen concentration in saliva samples from COVID-19 patients. Recently, a similar assay was successfully used to describe N concentration distributions in clinical NP swabs ^11^. Our study reveals that N concentrations in saliva span five orders of magnitude (0.2–1,000 pg/ml). Our assay displayed absolute specificity (in non-COVID-19 samples) and near-perfect sensitivity in correctly identifying samples with viral loads up to 35 CT cycles (by saliva PCR). Notably, the potential for infectivity has been shown even in cases with viral loads as low as ∼10,000 copies/ml (roughly corresponding to 33-34 CT cycles) ^15–17^. This highlights the need for ultrasensitive assays like the S-PLEX assay (LLD = 0.16 pg/ml) in order to minimize false negatives in salivary-based SARS-CoV-2 detection. As expected, the concordance of the three assays (S-PLEX saliva antigen, saliva-based PCR, paired NP swab-based PCR) was rather low in cases with very low viral loads (CT cycles ranging from 35–40), which reflects the borderline identification of these low viral load cases by all current methods. This result emphasizes the need for caution when interpreting method comparisons of COVID-19 assays, since no assay provides a perfect reference standard (especially for defining COVID-19 negativity).

There are some limitations in our study. First, we used a retrospectively collected cohort of frozen saliva samples collected at different time-points post symptom onset. Second, the size of our cohort was relatively small, especially for specificity evaluation where we only included 15 non-COVID-19 cases (as available via the existing REB). Ongoing efforts aim to validate this assay in a larger prospective study of COVID-19-patients with more comprehensive clinical annotation. Third, the cut-off for clinical positivity was arbitrarily set at 0.32 pg/ml (2 x LLD of the S-PLEX assay). Future studies with a larger cohort from non-COVID-19 and COVID-19 patients are needed to accurately define the best clinical cut-off for positivity of the S-PLEX assay.

This is the first proof-of-concept validation of the performance of the S-PLEX N assay in saliva-based SARS-CoV-2 detection. The ultrasensitivity and specificity of this assay and its applicability for saliva-based testing may render this test a valuable complementary alternative to PCR-based techniques, especially in cases where compliance to frequent swabbing may be questionable (e.g. schools and nursing homes). For mass screening, the S-PLEX N assay technology can complete 80 samples on one reader in about 3 hours and about 2,000 samples in 8 hours (by staggering plates), without any sample pretreatment, qualifying it as one of the most productive testing platforms for SARS-CoV2. Our preliminary finding presented here is the first step to unveiling a novel ultrasensitive approach that complements current PCR-based methods, which can help alleviate the analytical and operational challenges faced by mass SARS-CoV-2 testing strategies.

## Data Availability

The datasets used and/or analyzed during the current study are available from the corresponding author on reasonable request.

## References

1. Medicine, T. L. R. Realising the potential of SARS-CoV-2 vaccines—a long shot? Lancet Respir Medicine (2021) doi:10.1016/s2213-2600(21)00045-x.

2. Contreras, S. et al. The challenges of containing SARS-CoV-2 via test-trace-and-isolate. Nat Commun 12, 378 (2021).

3. Raffle, A. E., Pollock, A. M. & Harding-Edgar, L. Covid-19 mass testing programmes. Bmj 370, m3262 (2020).

4. Wyllie, A. L. et al. Saliva or Nasopharyngeal Swab Specimens for Detection of SARS-CoV-2. New Engl J Med 383, 1283–1286 (2020).

5. Silva, R. C. M. da et al. Saliva as a possible tool for the SARS-CoV-2 detection: a review. Travel Med Infect Di 38, 101920 (2020).

6. Comber, L. et al. Alternative clinical specimens for the detection of SARS□CoV□2: A rapid review. Rev Med Virol (2020) doi:10.1002/rmv.2185.

7. Michailidou, E., Poulopoulos, A. & Tzimagiorgis, G. Salivary diagnostics of the novel coronavirus SARS□CoV□2 (COVID□19). Oral Dis (2020) doi:10.1111/odi.13729.

8. Butler-Laporte, G. et al. Comparison of Saliva and Nasopharyngeal Swab Nucleic Acid Amplification Testing for Detection of SARS-CoV-2. Jama Intern Med 181, (2021).

9. Ning, B. et al. A smartphone-read ultrasensitive and quantitative saliva test for COVID-19. Sci Adv (2020) doi:10.1126/sciadv.abe3703.

10. Afzal, A. Molecular diagnostic technologies for COVID-19: Limitations and challenges. J Adv Res 26, 149–159 (2020).

11. Pollock, N. R. et al. Correlation of SARS-CoV-2 nucleocapsid antigen and RNA concentrations in nasopharyngeal samples from children and adults using an ultrasensitive and quantitative antigen assay. J Clin Microbiol (2021) doi:10.1128/jcm.03077-20.

12. Guglielmi, G. Fast coronavirus tests: what they can and can’t do. Nature 585, 496–498 (2020).

13. Jamal, A. J. et al. Sensitivity of nasopharyngeal swabs and saliva for the detection of severe acute respiratory syndrome coronavirus 2 (SARS-CoV-2). Clin Infect Dis ciaa848- (2020) doi:10.1093/cid/ciaa848.

14. Silva, J. et al. Saliva viral load is a dynamic unifying correlate of COVID-19 severity and mortality. Medrxiv Prepr Serv Heal Sci (2021) doi:10.1101/2021.01.04.21249236.

15. Singanayagam, A. et al. Duration of infectiousness and correlation with RT-PCR cycle threshold values in cases of COVID-19, England, January to May 2020. Eurosurveillance 25, 2001483 (2020).

16. Jaafar, R. et al. Correlation between 3790 qPCR positives samples and positive cell cultures including 1941 SARS-CoV-2 isolates. Clin Infect Dis ciaa1491- (2020) doi:10.1093/cid/ciaa1491.

17. Pekosz, A. et al. Antigen-based testing but not real-time PCR correlates with SARS-CoV-2 virus culture. undefined doi:10.1101/2020.10.02.20205708.

