## Supplemental Table 1 for "Ultrasensitive assay for saliva-based SARS-CoV-2 antigen detection"

**Running title:** Ultrasensitive saliva-based SARS-CoV-2 antigen detection

**Word count:** 2470

**Figures:** 2

**Tables:** 1

**Supplemental table:** 1

***To whom correspondence should be addressed**:

I. Prassas, Ph.D.

Mount Sinai Hospital, Joseph & Wolf Lebovic Ctr.,

60 Murray St [Box 32]; Flr 6 - Rm L6-201 Toronto, ON, M5T 3L9, Canada

and

E.P. Diamandis, Ph.D., M.D.

Mount Sinai Hospital, Joseph & Wolf Lebovic Ctr.,

60 Murray St [Box 32]; Flr 6 - Rm L6-201

Toronto, ON, M5T 3L9, Canada

**Supplemental Table 1 S-PLEX^®^ ultra-sensitive SARS-CoV-2 nucleocapsid antigen immunoassay and PCR test results for saliva samples from non-COVID-19 and COVID-19 cases**

|  |  |  | **Saliva sample** | | | |  | **Paired nasopharyngeal swab sample** | |
| --- | --- | --- | --- | --- | --- | --- | --- | --- | --- |
| Patient ID | Days since symptom onset | Days since diagnostic swab | PCR (Mean CT cycle) | PCR diagnosis (CT < 35 cut-off) | SARS-COV-2 N antigen concentration* (pg/ml) | N antigen test diagnosis** |  | PCR (Mean CT cycle) | PCR diagnosis (CT < 35 cut-off) |
| 1 | 12 | 5 | – | Negative | < 0.16 | Negative |  | 30 | Positive |
| 7 | 14 | 12 | – | Negative | < 0.16 | Negative |  | 30 | Positive |
| 10 | 14 | 4 | – | Negative | < 0.16 | Negative |  | 26 | Positive |
| 15 | 16 | 6 | 38 | Negative | < 0.16 | Negative |  | 29 | Positive |
| 21 | 26 | 5 | 35 | Negative | < 0.16 | Negative |  | 34 | Positive |
| 25 | 20 | 14 | 36 | Negative | < 0.16 | Negative |  | 39 | Negative |
| 26 | 11 | 6 | 35 | Negative | < 0.16 | Negative |  | – | Negative |
| 29 | 7 | 4 | – | Negative | < 0.16 | Negative |  | 27 | Positive |
| 33 | 27 | 14 | – | Negative | < 0.16 | Negative |  | – | Negative |
| 34 | 14 | 9 | – | Negative | < 0.16 | Negative |  | 35 | Positive |
| 39 | 15 | 14 | – | Negative | < 0.16 | Negative |  | – | Negative |
| 41 | 15 | 13 | – | Negative | < 0.16 | Negative |  | 34 | Positive |
| 43 | 15 | 14 | – | Negative | < 0.16 | Negative |  | 37 | Negative |
| 45 | 9 | 7 | – | Negative | < 0.16 | Negative |  | – | Negative |
| 54 | 10 | 10 | – | Negative | < 0.16 | Negative |  | 32 | Positive |
| 62 | 9 | 8 | – | Negative | < 0.16 | Negative |  | – | Negative |
| 63 | 13 | 12 | 36 | Negative | < 0.16 | Negative |  | – | Negative |
| 82 | – | – | – | Negative | < 0.16 | Negative |  | – | Negative |
| 83 | – | – | – | Negative | < 0.16 | Negative |  | – | Negative |
| 84 | – | – | – | Negative | < 0.16 | Negative |  | – | Negative |
| 85 | – | – | – | Negative | < 0.16 | Negative |  | – | Negative |
| 86 | – | – | – | Negative | < 0.16 | Negative |  | – | Negative |
| 87 | – | – | – | Negative | < 0.16 | Negative |  | – | Negative |
| 88 | – | – | – | Negative | < 0.16 | Negative |  | – | Negative |
| 89 | – | – | – | Negative | < 0.16 | Negative |  | – | Negative |
| 91 | – | – | – | Negative | < 0.16 | Negative |  | – | Negative |
| 92 | – | – | – | Negative | < 0.16 | Negative |  | – | Negative |
| 93 | – | – | – | Negative | < 0.16 | Negative |  | – | Negative |
| 94 | – | – | – | Negative | < 0.16 | Negative |  | – | Negative |
| 95 | – | – | – | Negative | < 0.16 | Negative |  | – | Negative |
| 96 | – | – | – | Negative | < 0.16 | Negative |  | – | Negative |
| 97 | – | – | – | Negative | < 0.16 | Negative |  | – | Negative |
| 98 | – | – | – | Negative | < 0.16 | Negative |  | – | Negative |
| 99 | – | – | – | Negative | < 0.16 | Negative |  | – | Negative |
| 100 | – | – | – | Negative | < 0.16 | Negative |  | – | Negative |
| 101 | – | – | – | Negative | < 0.16 | Negative |  | – | Negative |
| 102 | – | – | – | Negative | < 0.16 | Negative |  | – | Negative |
| 103 | – | – | – | Negative | < 0.16 | Negative |  | – | Negative |
| 104 | – | – | – | Negative | < 0.16 | Negative |  | – | Negative |
| 105 | – | – | – | Negative | < 0.16 | Negative |  | – | Negative |
| 81 | – | – | – | Negative | 0.17 | Negative |  | – | Negative |
| 51 | 11 | 9 | – | Negative | 0.17 | Negative |  | – | Negative |
| 56 | 28 | 27 | – | Negative | 0.18 | Negative |  | – | Negative |
| 75 | – | – | 31 | Positive | 0.18 | Negative |  | – | Positive |
| 71 | – | – | 33 | Positive | 0.19 | Negative |  | – | Positive |
| 20 | 15 | 4 | 33 | Positive | 0.20 | Negative |  | 33 | Positive |
| 46 | 11 | 9 | – | Negative | 0.20 | Negative |  | – | Negative |
| 17 | 16 | 10 | – | Negative | 0.20 | Negative |  | – | Negative |
| 4 | 8 | 3 | – | Negative | 0.21 | Negative |  | – | Negative |
| 42 | 15 | 9 | – | Negative | 0.24 | Negative |  | 27 | Positive |
| 8 | 9 | 9 | – | Negative | 0.25 | Negative |  | 29 | Positive |
| 3 | 14 | 11 | – | Negative | 0.26 | Negative |  | 25 | Positive |
| 48 | 7 | 7 | – | Negative | 0.27 | Negative |  | 36 | Negative |
| 6 | 3 | 2 | 30 | Positive | 0.29 | Negative |  | 36 | Negative |
| 65 | 12 | 10 | – | Negative | 0.30 | Negative |  | 32 | Positive |
| 52 | 21 | 8 | 36 | Negative | 0.31 | Negative |  | 18 | Positive |
| 38 | 5 | 3 | – | Negative | 0.36 | Weakly Positive |  | – | Negative |
| 14 | 7 | 5 | – | Negative | 0.37 | Weakly Positive |  | 29 | Positive |
| 50 | 22 | 17 | – | Negative | 0.39 | Weakly Positive |  | – | Negative |
| 40 | 19 | 17 | – | Negative | 0.41 | Weakly Positive |  | – | Negative |
| 9 | 10 | 5 | 34 | Positive | 0.44 | Weakly Positive |  | 33 | Positive |
| 37 | 33 | 15 | – | Negative | 0.45 | Weakly Positive |  | 37 | Negative |
| 58 | 16 | 4 | 37 | Negative | 0.45 | Weakly Positive |  | – | Negative |
| 5 | 4 | 4 | – | Negative | 0.48 | Weakly Positive |  | 28 | Positive |
| 24 | 16 | 14 | 30 | Positive | 0.51 | Weakly Positive |  | 34 | Positive |
| 23 | 13 | 9 | 31 | Positive | 0.52 | Weakly Positive |  | 22 | Positive |
| 59 | 10 | 6 | 34 | Positive | 0.55 | Weakly Positive |  | – | Negative |
| 22 | 8 | 5 | 34 | Positive | 0.58 | Weakly Positive |  | 25 | Positive |
| 28 | 11 | 3 | 37 | Negative | 0.61 | Weakly Positive |  | – | Negative |
| 31 | 14 | 3 | 28 | Positive | 0.67 | Weakly Positive |  | 27 | Positive |
| 57 | 4 | 2 | 33 | Positive | 0.72 | Weakly Positive |  | 27 | Positive |
| 30 | 14 | 3 | 36 | Negative | 0.79 | Weakly Positive |  | 24 | Positive |
| 73 | – | – | 32 | Positive | 0.98 | Weakly Positive |  | – | Positive |
| 66 | 4 | 4 | – | Negative | 0.99 | Weakly Positive |  | 28 | Positive |
| 27 | 11 | 5 | 29 | Positive | 1.1 | Weakly Positive |  | 33 | Positive |
| 16 | 18 | 14 | – | Negative | 1.4 | Weakly Positive |  | 30 | Positive |
| 72 | – | – | 28 | Positive | 1.5 | Weakly Positive |  | – | Positive |
| 90 | – | – | – | Negative | 1.6 | Weakly Positive |  | 22 | Positive |
| 11 | 13 | 7 | 32 | Positive | 1.8 | Weakly Positive |  | – | Negative |
| 80 | – | – | 31 | Positive | 4.5 | Strongly positive |  | – | Positive |
| 44 | 17 | 7 | 29 | Positive | 5.0 | Strongly positive |  | 28 | Positive |
| 60 | 8 | 8 | 29 | Positive | 5.2 | Strongly positive |  | 25 | Positive |
| 36 | 7 | 4 | 27 | Positive | 5.3 | Strongly positive |  | 21 | Positive |
| 2 | 11 | 8 | 31 | Positive | 5.9 | Strongly positive |  | 34 | Positive |
| 49 | 9 | 7 | 28 | Positive | 6.2 | Strongly positive |  | 24 | Positive |
| 61 | 6 | 4 | 33 | Positive | 6.2 | Strongly positive |  | 30 | Positive |
| 13 | 12 | 7 | 29 | Positive | 6.7 | Strongly positive |  | – | Negative |
| 78 | – | – | 27 | Positive | 8.2 | Strongly positive |  | – | Positive |
| 35 | 5 | 3 | 30 | Positive | 9.2 | Strongly positive |  | 15 | Positive |
| 53 | 25 | 11 | 26 | Positive | 12.0 | Strongly positive |  | – | Negative |
| 69 | 9 | 10 | 30 | Positive | 16.4 | Strongly positive |  | – | Positive |
| 77 | – | – | 24 | Positive | 26.9 | Strongly positive |  | – | Positive |
| 18 | 4 | 3 | 26 | Positive | 31.5 | Strongly positive |  | 34 | Positive |
| 47 | 42 | 8 | 23 | Positive | 32.4 | Strongly positive |  | – | Negative |
| 12 | 8 | 4 | 28 | Positive | 33.0 | Strongly positive |  | 21 | Positive |
| 64 | 24 | 18 | 25 | Positive | 53.3 | Strongly positive |  | 32 | Positive |
| 79 | – | – | 24 | Positive | 55.3 | Strongly positive |  | – | Positive |
| 32 | 7 | 1 | 24 | Positive | 70.9 | Strongly positive |  | 26 | Positive |
| 19 | 18 | 4 | 24 | Positive | 108.4 | Strongly positive |  | 31 | Positive |
| 76 | – | – | 24 | Positive | 128.4 | Strongly positive |  | – | Positive |
| 68 | 27 | 6 | 29 | Positive | 162.1 | Strongly positive |  | – | Positive |
| 70 | 1 | 1 | 27 | Positive | 544.5 | Strongly positive |  | 33 | Positive |
| 55 | 12 | 10 | 21 | Positive | > 1,000 | Strongly positive |  | 19 | Positive |
| 67 | 15 | 6 | 27 | Positive | > 1,000 | Strongly positive |  | – | Positive |
| 74 | – | – | 18 | Positive | > 1,000 | Strongly positive |  | – | Positive |

*Nucleocapsid (N) antigen concentration is reported as measured in the assay, without accounting for the sample dilution.

**Nucleocapsid (N) antigen test positivity cut-off (0.32 pg/ml) is set at the value of twice the lower limit of detection of the assay (0.16 pg/ml)

| Negative | Weakly positive | Strongly positive |
| --- | --- | --- |
